# Amyloid-associated increases in soluble tau is a key driver in accumulation of tau aggregates and cognitive decline in early Alzheimer

**DOI:** 10.1101/2022.01.07.22268767

**Authors:** Alexa Pichet Binette, Nicolai Franzmeier, Nicola Spotorno, Michael Ewers, Matthias Brendel, Davina Biel, ADNI, Olof Strandberg, Shorena Janelidze, Sebastian Palmqvist, Niklas Mattsson-Carlgren, Ruben Smith, Erik Stomrud, Rik Ossenkoppele, Oskar Hansson

## Abstract

For optimal design of anti-amyloid-β (Aβ) and anti-tau clinical trials, it is important to understand how Aβ and soluble phosphorylated tau (p-tau) relate to the accumulation of tau aggregates assessed with PET and subsequent cognitive decline across the Alzheimer’s disease (AD) continuum. In early stages of AD, increased concentration of soluble CSF p-tau was the main driver of accumulation of insoluble tau aggregates across the brain, and mediated the effect of Aβ on tau aggregation. Further, higher soluble p-tau concentrations were mainly related to faster accumulation of tau aggregates in the regions with strong functional connectivity to individual tau epicenters. In this early stage, higher soluble p-tau concentrations were associated with cognitive decline, which was mediated by faster increase of tau aggregates. In AD dementia, when Aβ fibrils and soluble p-tau levels have plateaued, cognitive decline was driven by the accumulation rate of insoluble tau aggregates. Our data suggest that therapeutic approaches reducing soluble p-tau levels might be most favorable in early AD.

## INTRODUCTION

Alzheimer’s disease (AD) is characterized by a cascade of molecular and neurodegenerative brain changes, in which Aβ plaques start to accumulate ∼20 years before symptom onset followed by the accumulation and spreading of neurofibrillary tau aggregates with ensuing neurodegeneration and AD dementia^1,2^. Multiple imaging studies of *in vivo* pathological processes with positron emission tomography (PET) in AD showed that cortical Aβ deposition precedes neocortical tau aggregation by several years^1,3^, and Aβ has been shown to amplify tau spreading in preclinical studies^4,5^. In contrast, age-related tau pathology typically occurring in the medial temporal lobe rarely spreads widely into the neocortex in the absence of cortical Aβ^6,7^, suggesting that the progression of tau aggregates is modified by Aβ^1,5^. Importantly, the spreading pattern of tau closely coincides with subsequent neurodegeneration^8^ and predicts future cognitive decline^9-11^, hence tau pathology shows strong correlations with clinical deterioration in AD^5,12^. However, it is still unclear by which mechanisms Aβ drives the spreading of tau pathology. Therefore, understanding the interplay between Aβ fibrils deposition and the accumulation and progression of tau aggregates during the different disease stages of AD is of high clinical relevance and will be crucial to identify potential targets for preventing the development of tau pathology and cognitive decline in AD.

Among the earliest tau-related abnormalities in AD patients are increases in soluble hyperphosphorylated tau (p-tau) concentrations, which can be detected in plasma and cerebrospinal fluid (CSF) during life^1^. Levels of p-tau start increasing already at the preclinical stage of AD when persons are asymptomatic^13,14^, increase even further in early symptomatic AD and reach a plateau in patients with AD dementia^15-17^. The rate of tau production^18^ and increases in soluble p-tau^19^ have been shown to correlate with Aβ burden, further strengthening the link between Aβ and tau. Soluble p-tau increases also precede the neocortical deposition of fibrillary tau pathology in AD^20-22^. Hence, we hypothesize that the Aβ-driven production and secretion of soluble p-tau may be a critical event during the pathogenesis of AD that facilitates accumulation of tau aggregates across the brain, which in turn might lead to cognitive decline.

A putative mechanism by which Aβ triggers tau secretion is via stimulation of neuronal activity ^23^. Extracellular Aβ has been consistently shown to induce neuronal hyperactivity^24,25^ and neuronal activity enhances tau secretion^26-28^. Therefore, a major pathway for the propagation of tau pathology in AD are synaptic connections, which are assumed to provide the roadmap for trans-neuronal tau spreading^29,30^. This is supported by preclinical work, showing that interstitial p-tau or injections of p-tau seeds can be taken-up by post-synaptic neurons and spread across anatomically connected rather than spatially adjacent brain regions^4,28,31,32^. We recently translated these preclinical findings to *in vivo* neuroimaging data by combining functional MRI-based connectomics with tau-PET in AD. We and others reported that connected brain regions show correlated tau accumulation rates and that patient-level tau spreading patterns follow the connectivity patterns of regions in which aggregated tau pathology emerges first (i.e, tau epicenters)^33-35^. Altogether, this would support a model in which i) Aβ leads to increased concentrations of soluble p-tau across the AD continuum, ii) greater concentrations of soluble p-tau are associated with faster formation of insoluble tau aggregates, and iii) abnormal tau spreads across connected neurons. Yet, such an integrative model of how Aβ fibrils, soluble p-tau concentrations, accumulation rates of insoluble tau aggregates and cognitive decline are interrelated across the entire AD clinical continuum is still missing. Here, we leveraged cross-sectional and longitudinal biomarker and cognitive data to test this model, and informed by the results we provide a working model proposing a pathway from the earliest pathological beginnings towards the clinical syndrome of AD dementia. We first tested how soluble p-tau concentrations measured in CSF and local Aβ fibrils affected the local accumulation rate of tau aggregates in the same brain regions. Second, we investigated if soluble p-tau is a driver of connectivity-based accumulation of tau aggregates across the brain by combining CSF p-tau, longitudinal tau-PET and resting-state functional connectivity. Lastly, we aimed to elucidate the mechanisms that lead to cognitive decline across the whole AD continuum, bringing together both measures of soluble p-tau and accumulation of aggregated tau as they follow more closely the apparition of symptoms than Aβ pathology ^9^.

## RESULTS

### Study design and participants

The study was conducted in one of the largest samples available worldwide with cross-sectional Aβ-PET and soluble CSF p-tau measurement, and longitudinal tau-PET and cognition, covering the full spectrum of AD. The Swedish BioFINDER-2 cohort was the primary cohort of interest, from which participants with all abovementioned measures were included. The final sample included 327 participants composed of 160 cognitively unimpaired (CU) Aβ-negative participants, 51 Aβ-positive CU, 54 Aβ-positive patients with mild cognitive impairment (MCI) and 62 Aβ-positive individuals with AD dementia (Table 1). The CU and MCI Aβ-positive participants were grouped together to study the early stages of AD, hereafter referred to as “non-demented participants”. To quantify the regional levels of insoluble tau aggregates, tau-PET was performed using the second-generation radiotracer [^18^F]RO948, shown to be sensitive to early tau aggregates^36,37^. Insoluble Aβ aggregates were measured with Aβ-PET using [^18^F]flutemetamol as the radiotracer. All PET data were parcellated in 200 cortical regions-of-interest using the Schaefer brain atlas^38^. Concentrations of soluble p-tau were measured using p-tau217 in the CSF^39^. The main results focusing on early AD were also validated in the Alzheimer’s Disease Neuroimaging Initiative (ADNI) database using [^18^F]flortaucipir PET, CSF p-tau181 as well as [^18^F]florbetapir or [^18^F]florbetaben Aβ-PET (n=119: 52 CU Aβ-negative, 36 Aβ-positive CU and 31 Aβ-positive MCI participants; Extended Data Table and Fig. 1). Given the smaller sample size in ADNI, only analyses where results could be detected with sufficient power calculated based on the BioFINDER-2 analyses were conducted. In addition, associations including regional Aβ-PET were not tested since centiloid assessment for regional Aβ SUVR across different tracers is not yet established. All results pertaining to the ADNI cohort are described in the on-line Extended Data.

**Table 1.** BioFINDER-2 cohort demographics. Data are presented as mean ± standard deviation unless specified otherwise. One Aβ-positive non-demented participant had missing *APOE* genotype. mPACC scores are z-scores. Abbreviations: Aβ= beta-amyloid; *APOE*ε4= apolipoprotein E genotype (carrying at least one ε4 allele); CSF p-tau217= cerebrospinal fluid phosphorylated tau 217; MMSE= Mini-Mental State Examination; mPACC= modified preclinical Alzheimer cognitive composite; PET= positron emission tomography.

| | Aβ-negative controls<br>(n=160) | Aβ-positive non-demented<br>(n=105) | Aβ-positive AD dementia<br>(n=62) |
| --- | --- | --- | --- |
| **Age** (years) | 62.4 ± 13.7 | 70.6 ± 8.1 | 72.8 ± 7.1 |
| **Sex F** (%F) | 73 (46%) | 49 (47%) | 35 (56%) |
| **Education** (years) | 12.9 ± 3.3 | 12.3 ± 3.8 | 12.0 ± 4.7 |
| **APOEε4 carriers** (%) | 60 (38%) | 79 (76%) | 41 (66%) |
| **CSF p-tau217** (pg/ml) | 46.9 ± 31.1 | 242.4 ± 171.0 | 570.7 ± 306.5 |
| **MMSE** | 29.0 ± 1.2 | 27.7 ± 1.9 | 19.8 ± 4.3 |
| **mPACC** | 0.01 ± 0.7 | -1.3 ± 1.2 | -4.4 ± 1.5 |
| **tau-PET follow-up time** (years) | 1.8 ± 0.2 | 1.8 ± 0.2 | 1.6 ± 0.3 |

**Figure 1.**
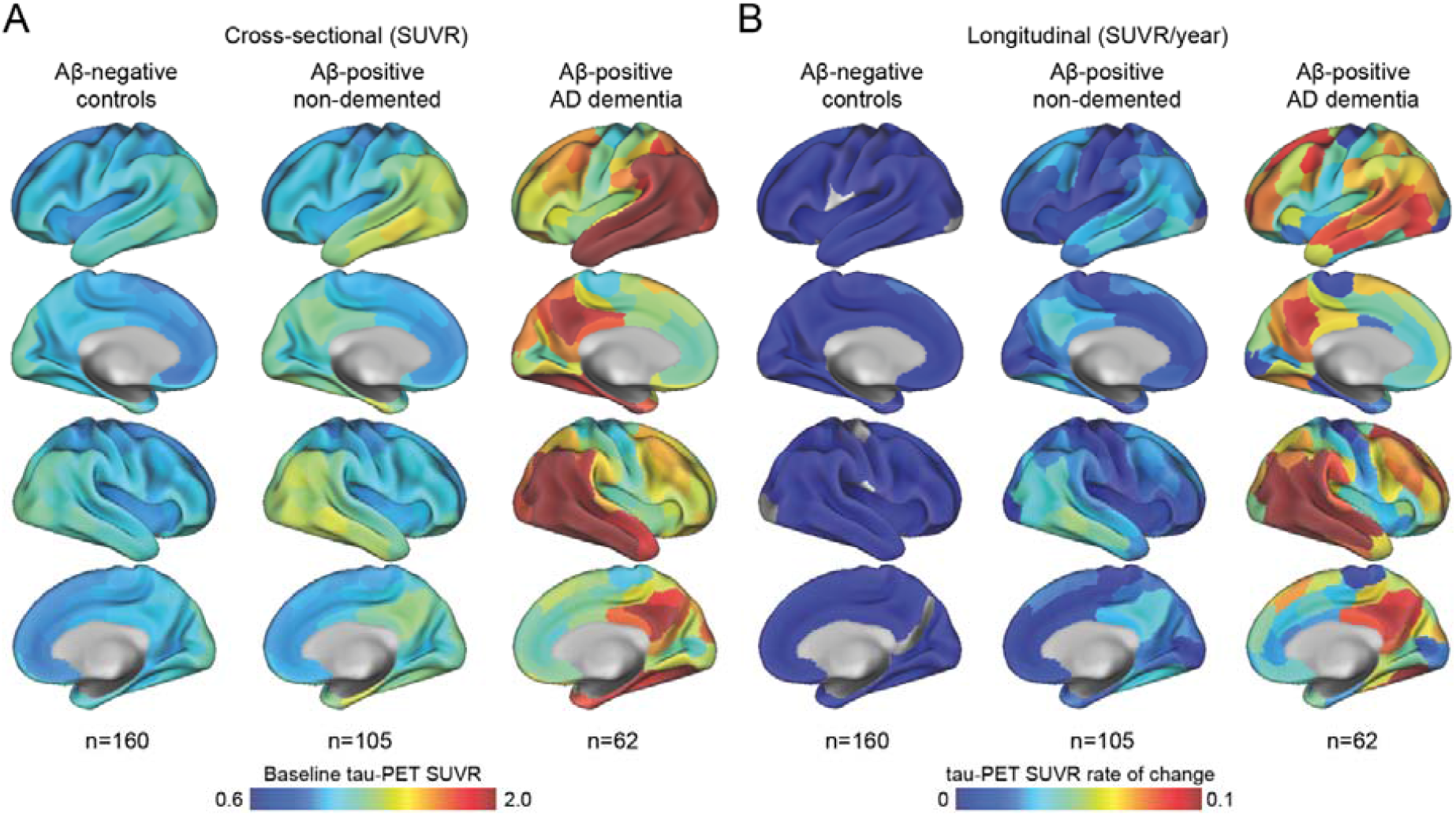
Mean spatial distribution of cross-sectional tau-PET [^18^F]RO948 SUVR and longitudinal rate of change. (A) Surface renderings of average baseline tau-PET SUVR in Aβ-negative controls, Aβ-positive non-demented participants and Aβ-positive patients with AD dementia in the 200 parcels from the Schaefer 200-ROI atlas (B) Surface renderings of yearly tau-PET SUVR rate of change derived as the slope from linear mixed-effect models in the same participants group as in (A)

In the BioFINDER-2 cohort, the baseline distribution of insoluble tau aggregates assessed via tau-PET recapitulated the AD-typical deposition in the medial and lateral temporal lobes early on, before gradually extending mainly into lateral and medial parietal and lateral occipital regions at symptomatic AD stages (Fig. 1A). The longitudinal accumulation rate of tau aggregates (i.e., the annual tau-PET rate of change measured over a mean time of 1.8 years) was highest in temporal lobe regions in non-demented participants, and showed more extensive involvement mainly of medial parietal and lateral frontal regions in AD dementia (Fig. 1B). Quantitatively, in regions-of-interest approximating Braak I to IV (encompassing medial and lateral parts of the temporal lobe), the average annual SUVR rate of change was 0.8% in Aβ-negative controls, 2.4% in Aβ-positive non-demented individuals, and 5.7% in Aβ-positive patients with AD dementia. In comparison, focusing on the top 10% regions with the highest accumulation rate in the dementia stage, which encompasses mainly lateral temporal and parietal regions, AD patients had an average accumulation of tau aggregates of 10.5% per year, and Aβ-positive non-demented individuals, of 3.6%. The rate of change remained unchanged at 0.8% in Aβ-negative controls.

### Increased soluble p-tau is the main modifier of tau aggregate accumulation rates in early AD

First, we tested whether either the level of local Aβ aggregates and/or the concentrations of soluble p-tau were most strongly associated with local increases in insoluble tau aggregates over time in early stages of AD using linear regression models in each of the 200 brain regions. Across Aβ-positive non-demented participants, regional Aβ was positively associated with greater accumulation of tau aggregates over time, most prominently in temporo-parietal regions (Fig. 2A). On average across the 118 regions surviving correction for multiple comparisons, the standardized estimate of Aβ was 0.29 (range 0.21 to 0.44, average p-values in those regions= 0.02). Levels of soluble p-tau was also positively correlated with accumulation of tau aggregates, but in contrast to regional Aβ this was observed in nearly all brain regions (n=189) and the effect sizes were substantially higher (Fig. 2A, average standardized estimate: 0.45, range 0.21 to 0.76, average p-values <0.001). When using regional Aβ and soluble p-tau simultaneously as predictors (Fig. 3B), the widespread CSF p-tau associated tau accumulation pattern remained virtually the same. In contrast, for regional Aβ there where almost no regions that remained significant after accounting for soluble p-tau. Furthermore, all key regions where soluble p-tau was most strongly associated with accumulation of tau aggregates over time remained significant even when additionally accounting for baseline levels of tau aggregates in each region (Fig. 3C), albeit with slightly lower estimates (average standardized estimate: 0.25, range: 0.11 to 0.45, average p-values= 0.01).

**Figure 2.**
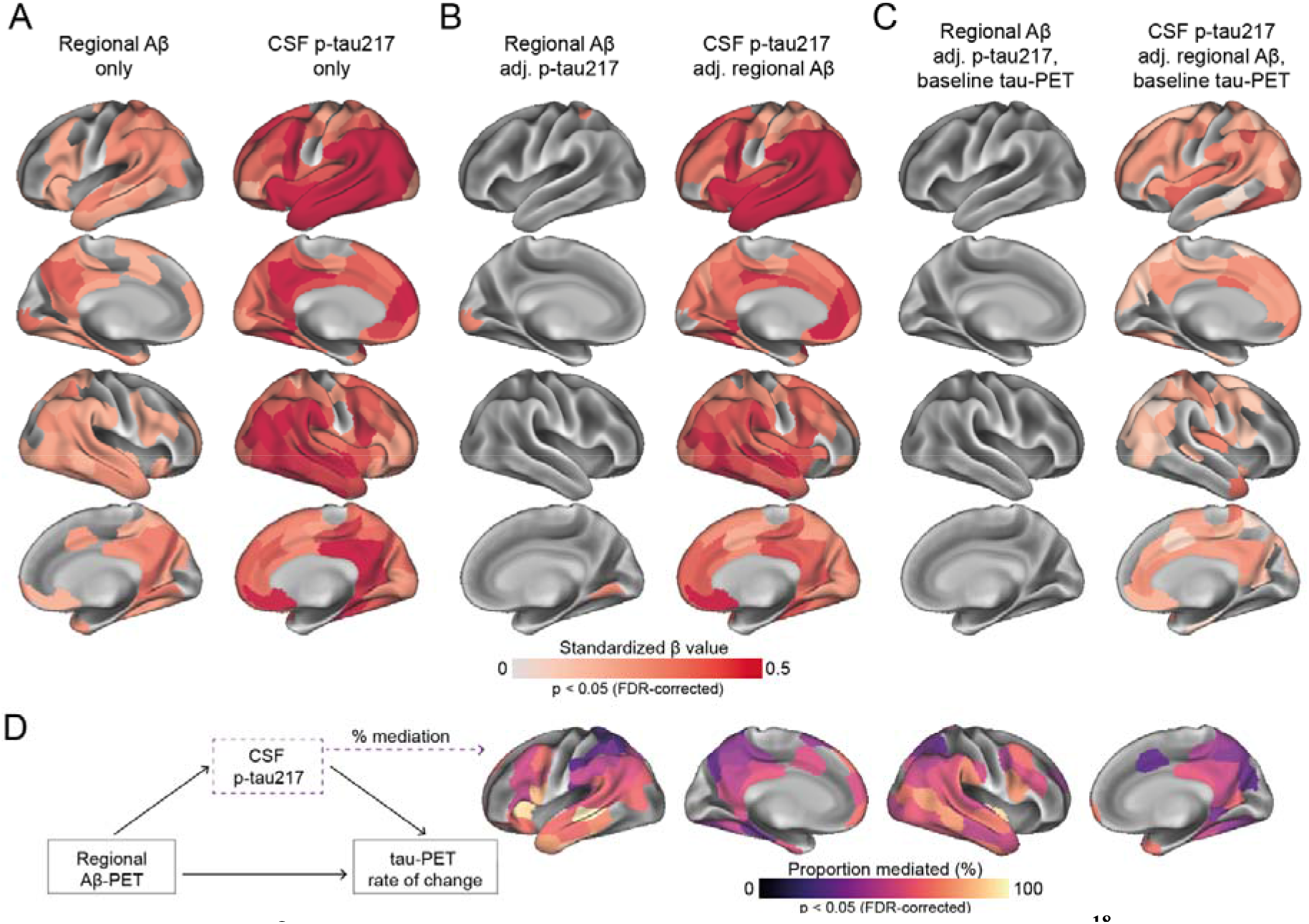
Regional Aβ-PET and CSF p-tau217 associations with regional tau-PET [^18^F]RO948 accumulation in Aβ-positive non-demented participants. (A) Standardized beta coefficient of local Aβ-PET in regions where regional Aβ-PET flutemetamol SUVR (left column) relates to regional tau-PET rate of change, adjusting for age and sex. Right columns were derived from a similar model, but using CSF p-tau217 as predictor instead of Aβ-PET (B) Standardized beta coefficient of local Aβ-PET (left column) and CSF p-tau217 (right column) is associated to regional tau-PET rate of change when including both biomarkers in the same model, adjusting for age and sex (tau PET rate of change ∼ regional Aβ-PET + CSF p-tau217 + age + sex) (C) Same depiction as in (B), when additionally controlling for regional baseline tau-PET SUVR (D) Mediating effect of CSF p-tau217 on local Aβ-related accumulation of local tau aggregates. The mediation models were performed region-wise, and the percentage of the mediating effect are projected on the brains. All regions shown on the brain are significant at p<0.05 after FDR-correction. See Extended Data Fig. 2 for similar results in ADNI.

**Figure 3.**
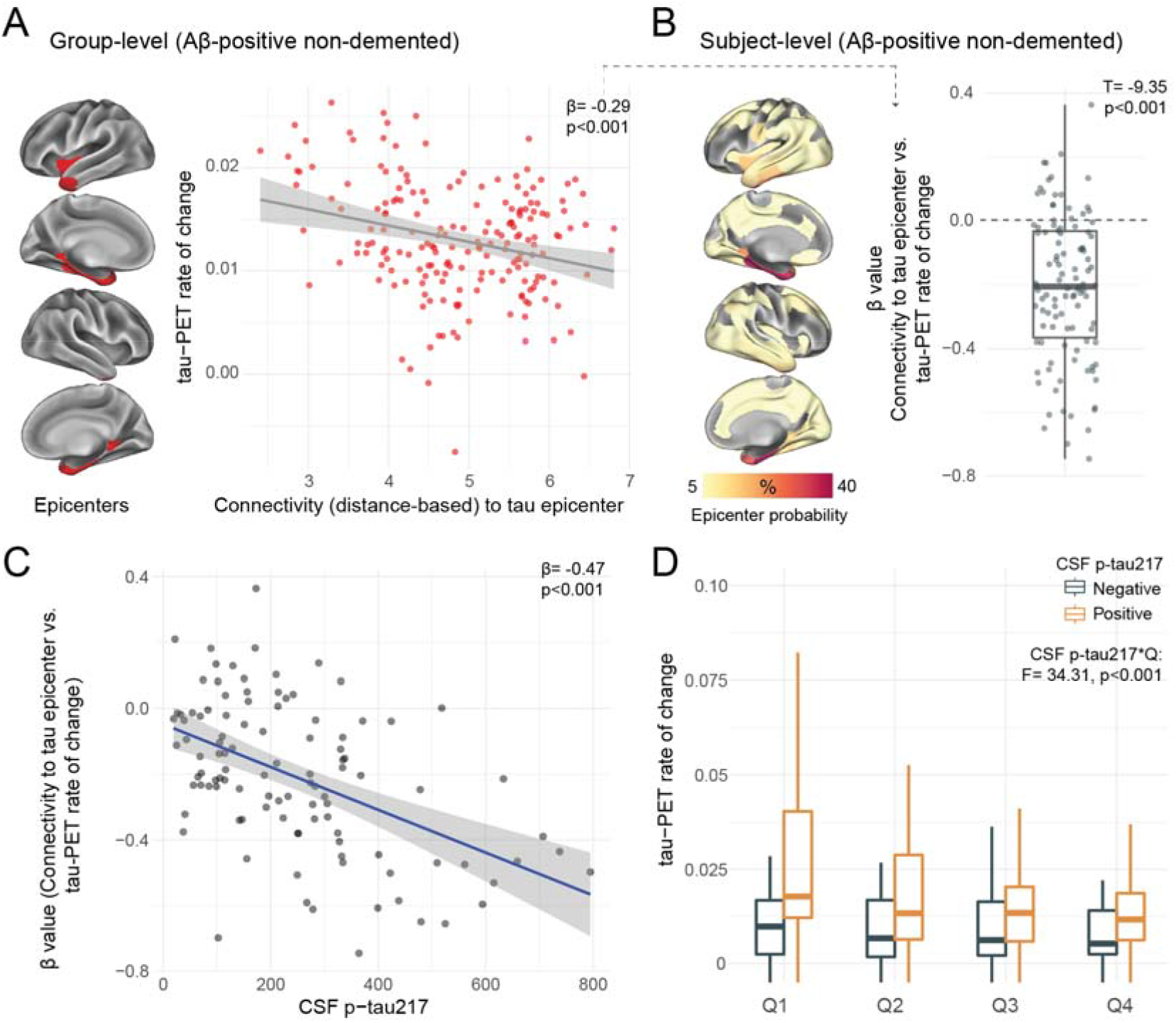
Individualized connectivity-based associations of tau-PET rate of change over time and CSF p-tau217 in Aβ-positive non-demented participants. (A) Group-level analysis showing how connectivity to the tau epicenters (projected on the glass brains) relates to tau-PET rate of change across the whole brain. Each dot represents a brain region. Regions more strongly functionally connected to the epicenters have greater rate of tau-PET accumulation (B) Repeating the same approach depicted in (A) at the individual level, the values on the glass brains represent the percentage that each region is classified as an epicenter. The box plot shows the individual β-value from the correlation between tau-PET rate of change and connectivity-based distance to epicenters across all brain regions (C) Scatter plot of the associations between CSF p-tau217 and the β-values of epicenter connectivity to tau-PET rate of change. The expected negative association suggests that higher CSF p-tau217 is associated with the overall pattern of tau-PET change in more functionally connected regions to epicenters (D) Average tau-PET rate of change in regions split into quartiles based each region’s connectivity to the tau epicenters (Q1 represents top 25% regions with strongest functional connectivity to the epicenters, etc.). See Extended Data Fig. 3 for similar results in ADNI.

Lastly, based on the observed interplay between local Aβ and soluble p-tau on the increased rate of tau aggregates accumulation, we formally tested the mediating effect of soluble p-tau on Aβ-related tau-PET rate of change at the regional level. Fig. 2D summarizes the proportion of mediating effect of soluble p-tau on all regions surviving multiple comparisons (n=108), which averaged to 54% across all regions. Overall, the results indicate that the effects of local Aβ aggregates on the accumulation rate of insoluble tau aggregates over time is largely mediated by increased concentrations of soluble p-tau.

### Soluble p-tau levels relate to connectivity-based accumulation of tau aggregates in early AD

Another important modifier of regional tau accumulation is the functional architecture of the brain. We therefore investigated whether the association between connectivity-based accumulation of tau aggregates was influenced by the concentration of soluble p-tau. Briefly, after defining participant-specific tau-PET epicenters (i.e., the top 10 regions with the highest tau-PET SUVR probability from Gaussian-mixture modeling at baseline), accumulation of tau aggregates in the remaining 190 regions was correlated to functional connectivity strength to the epicenters^33,40^. We first applied the model at the group-level, where we found greater accumulation rates of insoluble tau aggregates in regions more strongly connected to the epicenters (shorter distance-based connectivity) in early AD (Fig. 3A). Next, we applied this model at the individual level. We found that higher accumulation rates of tau aggregates were observed in regions that showed strongest connectivity to the tau epicenters defined on baseline tau-PET, as evidenced by negative β-values (i.e. reflecting the association between connectivity to the epicenters and tau aggregates accumulation across all non-epicenter ROIs; Fig. 3B)

Next, given the demonstrated association between soluble p-tau and the accumulation of tau aggregates, we hypothesized that higher soluble p-tau concentrations would relate to a stronger association between connectivity to the epicenters and the accumulation rates of tau aggregates in the remaining brain regions. Across all non-demented Aβ-positive participants, we found that participants with higher soluble p-tau levels had a stronger association between connectivity to epicenter and tau-PET rate of change in non-epicenter ROIs (β-value), while accounting for global Aβ, age and sex (Fig. 3C). Analyzing the data in a complementary way, we measured the rate of tau aggregates accumulation in regions split into quartiles, i.e. average tau-PET rate of change in individualized top 25% regions with the greatest connectivity to tau epicenters as quartile 1 (Q1), up to quartile 4 (Q4, regions with the lowest connectivity to tau epicenters). Repeated measures ANOVA revealed an interaction of soluble p-tau levels and quartiles (F=34.3, p<0.001), suggesting the importance of soluble p-tau and connectivity-based regions on the accumulation rates of insoluble tau aggregates (Figure 3D). This effect was particularly evident Q1, which showed the greatest effect size between CSF p-tau-positive and p-tau-negative participants (Cohen’s d=0.70 vs. 0.54 to 0.60 in Q2 to Q4).

### Accumulation of insoluble tau aggregates mediates the associations between soluble p-tau and cognitive decline in early AD

With soluble p-tau being related to local accumulation of tau aggregates and connectivity-mediated tau accumulation, we then investigated how those different tau measures related to cognitive decline in early stages of AD (i.e. Aβ-positive non-demented participants). Here we focused on tau-PET rate of change in regions most strongly connected to the epicenters (i.e. Q1), to capture individualized level of tau aggregates accumulation. All different tau-related measures (CSF p-tau217, tau-PET rate of change in Q1 and the β-value of connectivity-based tau-PET rate of change) were associated with cognitive decline as measured by annual change on the modified preclinical Alzheimer’s cognitive composite (mPACC) scores over time, designed to capture cognitive changes in the earliest AD stages (Fig. 4A-C). Given the observation that soluble p-tau increases early in AD and prior to the formation of neocortical insoluble tau aggregates, we then tested whether measures related to the accumulation rate of insoluble tau aggregates mediated the association between soluble p-tau concentrations and subsequent cognitive decline. First, the strength (β-value) of the association between accumulation rate of tau aggregates and functional connectivity to tau epicenters mediated 41% of the association between soluble p-tau and cognitive decline (Fig. 4F), suggesting a mediating effect of faster connectivity-based tau aggregates accumulation. Second, the accumulation rate of tau aggregates in the regions most connected to the subject-specific tau epicenter (Q1) mediated 46% of the association between soluble p-tau concentrations and the rate of cognitive decline (Fig. 4G). In sensitivity analyses, using the slopes of MMSE scores (instead of mPACC slopes) to measure cognitive decline, we found very similar mediating effects of tau aggregation (for mediating effects of the connectivity-based β-value: c’-c= −0.16 [95% CI −0.32, −0.02], p=0.02; for mediating effect of the tau-PET rate of change in Q1: c’-c= −0.18 [95% CI −0.32, −0.04], p=0.02). Taken together, the results indicate that higher concentrations of soluble p-tau are associated with cognitive decline in early AD, which is partly mediated by increased accumulation rates of insoluble tau aggregates.

**Figure 4.**
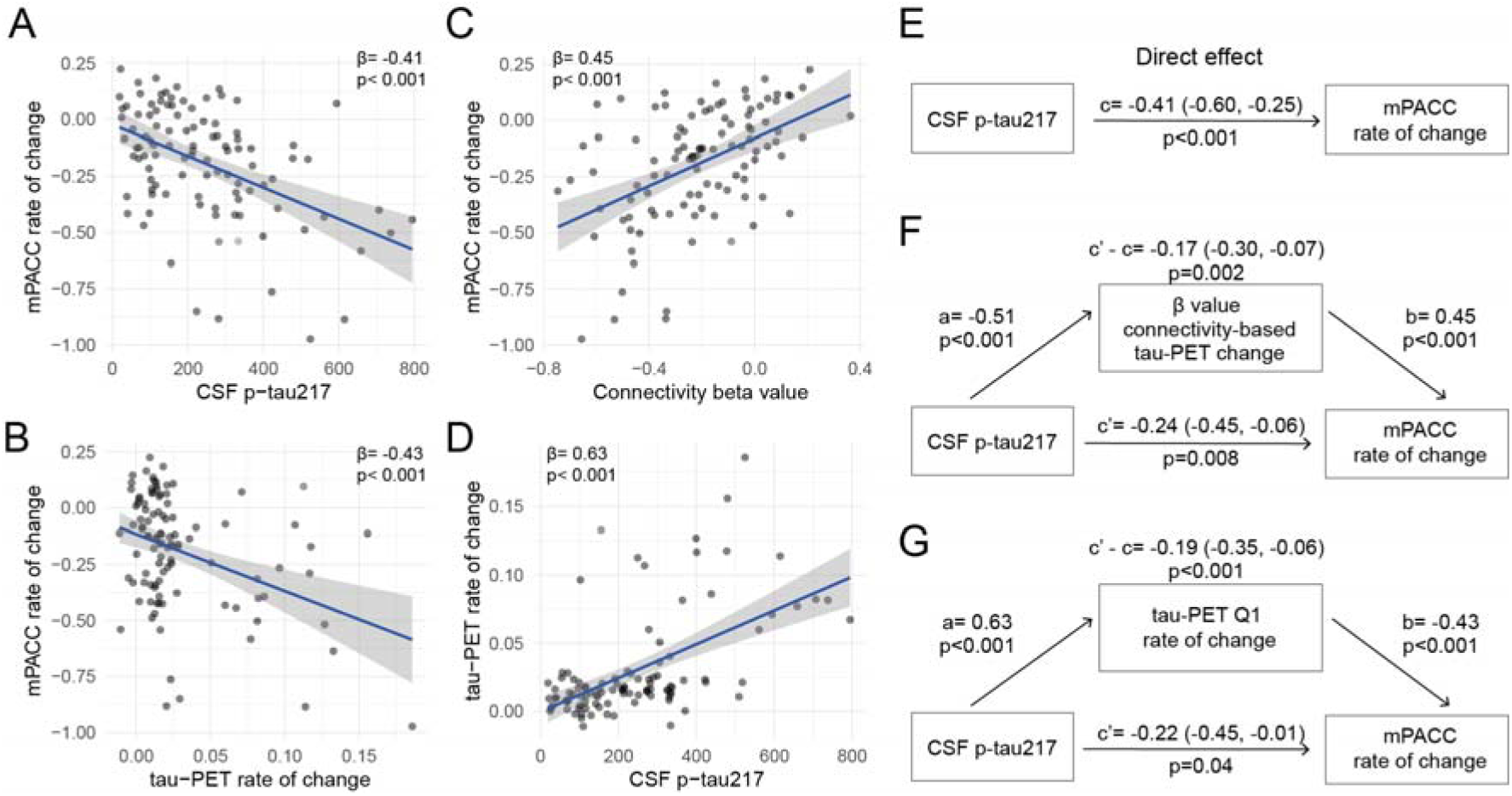
Tau aggregates accumulation mediates associations between CSF p-tau217 and cognitive decline in Aβ-positive non-demented participants. (A-D) Scatter plots of associations relevant to subsequent mediation analyses, beta coefficients from linear regressions adjusting for age, sex and education are reported (A) Association between CSF p-tau217 and mPACC rate of change (B) Association between tau-PET rate of change in Q1 and mPACC rate of change (C) Association between β-value from the correlation between tau-PET rate of change and connectivity-based distance to epicenters across all brain regions and mPACC rate of change (D) Association between CSF p-tau217 and tau-PET rate of change in Q1 (E-G) Mediation analysis of the relationship between CSF p-tau217, measures of tau-PET and cognitive decline measured as the rate of change on mPACC. The direct effect (c) of CSF p-tau217 on cognitive decline is shown in (E). Analyses are shown with β-value based on connectivity and tau-PET change (F), and tau-PET rate of change in Q1 (G) as mediators. The mediated effect is designated *c*-*c*′. The remaining effect of CSF p-tau217 on cognitive decline after adjusting for the mediator is designated *c*′. 95% confidence intervals derived from 1000 simulations are reported in parentheses. The direct effect of CSF p-tau217 on the mediator is *a*, and the direct effect of the mediator on cognitive decline is *b*. The β-value based on connectivity and tau-PET change (F) and tau-PET rate of change (G) partially mediated the relationship between CSF p-tau217 and cognitive decline. To facilitate model comparisons, all models use continuous standardized (*z-*score) data for variables of interest.

### Distinct associations between soluble p-tau and accumulation of tau aggregates in AD dementia

All previous analyses focused on non-demented participants, to study the effects of soluble p-tau and connectivity on accumulation of tau aggregates in early stages of AD. To investigate the full AD clinical continuum, we repeated the main analyses focusing on Aβ-positive individuals with AD dementia. Our motivation to focus on AD dementia patients separately was based on the plateau-phase reached by soluble p-tau species in more advanced disease stages (Fig. 5A-B). In Aβ-positive non-demented participants, higher soluble p-tau concentrations were associated with faster tau aggregates accumulation, above levels of Aβ and baseline tau aggregates. In contrast, in AD dementia, soluble p-tau concentrations were not associated with the accumulation rates of tau aggregates in any region after adjusting for baseline levels of local tau aggregates (Fig. 5C). The expected negative association between greater regional accumulation rates of tau aggregates and connectivity to the tau epicenters across the brain was present at the dementia stage, but the strength of this association (β-value) was not related to soluble p-tau levels (Fig. 5D). Similarly, soluble p-tau concentrations did not have a direct effect on cognitive decline (Fig. 5E). Rather, the accumulation rate of tau aggregates over time was most associated with cognitive decline at this stage of the disease (Fig. 5F). Further, soluble Aβ levels, measured as the CSF ratio of Aβ42/40, were not related to the accumulation of tau aggregates (β= 0.10, p=0.37 for tau-PET rate of change in Q1) or cognitive decline (β= −0.25, p=0.08 for MMSE slope). Overall, these results suggest that the accumulation rate of tau aggregates and cognitive decline seem independent of soluble p-tau concentrations in the dementia stage of the disease.

**Figure 5.**
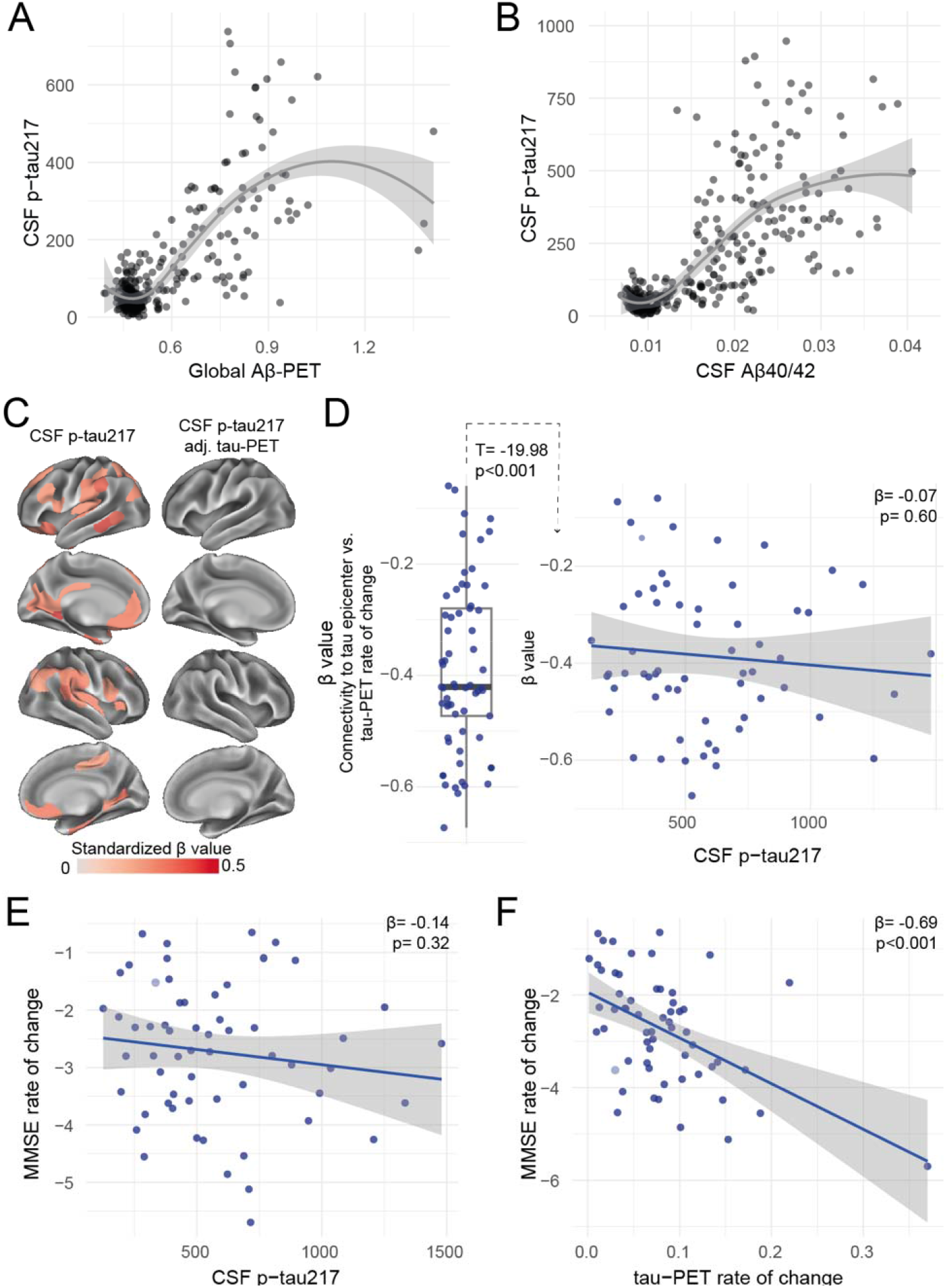
Distinct associations between CSF p-tau217 and tau-PET rate of change in the AD dementia stage. (A) Associations between global Aβ-PET SUVR (referenced to pons) from flutemetamol and CSF p-tau217. Note that AD dementia patients within BioFINDER-2 undergo lumbar punctures but not Aβ-PET, explaining the gap in highest values. (B) Associations between the ratio of CSF Aβ40/42 from Elecsys and CSF p-tau217. Both associations (in A-B) are nonlinear and flatten at high Aβ load (C) CSF p-tau217 alone was mildly associated with tau-PET rate of change (left column), which was not the case when additionally adjusting for regional baseline tau-PET SUVR (right column) (D) Box plot showing the expected negative β-value from the correlation between tau-PET rate of change and connectivity-based distance to epicenters across all AD patients, but this β-value was not related to CSF p-tau217 levels (E-F) Scatter plots of associations between CSF p-tau217 (E) and tau-PET rate of change in Q1 (F) and cognitive decline, as measured by MMSE rate of change. Beta coefficients from linear regressions adjusting for age, sex and education are reported. The rate of tau aggregates accumulation was related to cognitive decline, which was not the case for CSF p-tau217.

## DISCUSSION

The major aim of the present longitudinal biomarker study was to assess the relations between soluble p-tau concentrations and Aβ-related accumulation of insoluble tau aggregates over time in AD. In a large sample of Aβ-positive non-demented individuals of the BioFINDER-2 cohort, we found first, that increases in soluble CSF p-tau drove the association between local Aβ fibrils and the subsequent accumulation of tau aggregates measured with PET and second, that elevated soluble p-tau was associated with faster accumulation of tau aggregates across functionally connected brain regions. Importantly, these findings were fully replicated in the ADNI cohort. Third, we found that elevated soluble p-tau concentrations were associated with faster cognitive decline in early stages of AD, which was mediated by faster accumulation rates of tau aggregates. However, soluble p-tau concentrations, that plateau at the late-stage of Aβ accumulation, were no longer associated with local accumulation of tau aggregates or cognitive decline in patients with AD dementia. At this advanced stage, baseline levels of tau aggregates predicted subsequent accumulation of tau aggregates in the same brain regions, which was related to greater cognitive decline. These results suggest that self-replication of tau aggregates may drive tau accumulation and clinical deterioration once Aβ and p-tau levels have plateaued in late-stage AD. Taken together, these findings are congruent with the hypothesis that Aβ-induced soluble p-tau concentrations play a key role in initiating the aggregation and connectivity-mediated accumulation of tau pathology in early-stage AD and that local tau seeding and self-replication predominate once soluble p-tau concentrations have plateaued in AD dementia. This might have implications for clinical trials, since drugs reducing soluble p-tau concentrations (like anti-Aβ therapies or genetic anti-tau treatments) may be promising therapeutic strategies to prevent further accumulation and spread of tau aggregates and cognitive decline in early stages of AD. On the other hand, during the dementia stage of AD, directly targeting the local tau aggregates might prove to be more adequate than targeting soluble p-tau species or Aβ.

We found that soluble p-tau is a key driver of the Aβ-related accumulation of tau aggregates over time in early AD. Higher levels of CSF p-tau were strongly associated with longitudinal increases in tau aggregates in AD-specific temporo-parietal brain regions. Importantly, these effects survived adjustment for baseline levels of Aβ and tau aggregates in the same brain region, suggesting a unique contribution of CSF p-tau to the accumulation of tau aggregates. In addition to the previously demonstrated importance of baseline tau-PET levels for future tau aggregates accumulation^41,42^, we further showed the importance of soluble p-tau in this tau aggregation pathway. Moreover, soluble p-tau mediated the association between Aβ and accumulation rate of subsequent tau aggregates, building on previous cross-sectional results^19,21^. As such, Aβ-related increases in soluble p-tau may be a key initial step in the Aβ cascade that determines the accumulation of aggregated tau pathology. This is supported by previous cross-sectional studies, showing that increases in different CSF and plasma p-tau species (e.g. p-tau181, p-tau217, p-tau231) are among the first tau-related biomarker abnormalities across the AD continuum and are closely related to Aβ deposition^17,43-45^. Importantly, higher levels of soluble p-tau were also related to greater cognitive decline over time in Aβ-positive non-demented individuals, which was mediated by accelerated connectivity-based tau spreading and faster tau aggregates accumulation. In other words, increased concentrations of p-tau might drive the initial expansion and accumulation of tau aggregates, thereby leading to faster cognitive decline in early AD. This finding expands on previous studies linking longitudinal tau-PET and cognitive decline^46,47^, by further highlighting the importance of soluble p-tau in early stages of the disease.

From a pathophysiological point of view, Aβ has been shown to trigger increased synthesis, hyperphosphorylation and secretion of tau proteins from neurons, which leads to elevated interstitial p-tau concentrations, that pass into CSF and blood plasma where it can be detected *in vivo* using biomarkers^*1,18*^. Preclinical studies provide strong support for neuronal activity as a putative link between Aβ and tau secretion and accumulation. Aβ has consistently been shown to induce a hyper-excitatory shift in neuronal activity^24,48,49^, and this neuronal hyperexcitability may then induce increased p-tau secretion and subsequent spreading, with neuronal tau secretion being drastically enhanced by elevated neuronal activity^23,26,27^. This view is supported by our result showing that elevated soluble p-tau was associated with faster accumulation of tau aggregates across functionally connected brain regions. While accumulation of tau aggregates in a given brain region was related to its functional connectivity to tau epicenters^33,40,50^, the accumulation of tau aggregates from local epicenters to connected regions was enhanced in the face of abnormal soluble p-tau. Overall, both connectivity and soluble p-tau were important factors of the rate of tau aggregates accumulation. These findings suggest that Aβ-related soluble p-tau increases are a key prerequisite for the expansion of tau aggregates across functionally connected brain regions in early AD. A potential mechanism could be an Aβ-related increase of interstitial p-tau seeds that leads to increased uptake by post synaptic neurons, with ensuing local template-based misfolding and aggregation of physiological tau^28,51^.

Importantly, the results described above only apply to early pre-dementia AD stages and not to the dementia stage of AD. We found that the key role of soluble p-tau in the accumulation process of tau aggregates was no longer observed in patients with AD dementia. This can likely be explained by a plateau stage of Aβ aggregates and soluble p-tau levels occurring in more advanced disease stages^19,52^. In our data, soluble p-tau levels had a minor effect on further accumulation of tau aggregates and connectivity-based tau spreading in AD dementia, and only the tau aggregate accumulation rate was associated with cognitive decline (and not p-tau levels).

The main strength of this study is the integration of cross-sectional and longitudinal fluid and neuroimaging biomarkers of AD pathology and longitudinal cognitive measures across the clinical spectrum of AD. There are also several limitations. First, although we are expanding on previous studies that were limited by shorter follow-up or smaller sample sizes^53-55^, a longer follow-up time of tau-PET would increase the rate of tau aggregates accumulation in earlier stages of AD. Second, there were some differences between the biomarkers and cognitive data between BioFINDER-2 and ADNI. In BioFINDER-2 the focus was on CSF p-tau217, having shown greatest association with Aβ^15,56^, but the main results could still be validated in ADNI where only CSF p-tau181 was available. The two tau radiotracers used in the study present different unspecific binding, being mainly around the skull and meninges with [^18^F]RO948^57,58^ (used in BioFINDER-2) and in the choroid plexus (anatomically proximate to the hippocampus) and basal ganglia with flortaucipir (used in ADNI). To circumvent that unspecific binding would have affected our results, the brain parcellation chosen did not include the hippocampus or subcortical structures, and careful quality control was done to exclude participants with high meningeal binding on [^18^F]RO948. Unfortunately, cognitive decline could not be investigated in ADNI due to the small number of participants with longitudinal cognitive data, but we showed the robustness of these results in BioFINDER-2 by using two cognitive scores. Also, since AD patients did not undergo Aβ-PET in BioFINDER-2, we used the CSF Aβ42-40 ratio as an alternative biomarker of Aβ pathology.

In conclusion, reconciling both early and later stages of the disease, we propose an integrative model of how Aβ fibrils, soluble p-tau concentrations, functional connectivity, accumulation rates of insoluble tau aggregates and cognitive decline are interrelated across the entire clinical continuum of AD (Fig. 6). In this model, initially the Aβ-related increase of p-tau seeds are taken up by neurons to initiate the misfolding and aggregation of tau in a given brain region, with more substrate (i.e. soluble p-tau) for aggregation leading to more rapid local tau aggregation. Later on, local self-replication and accumulation of misfolded tau aggregates may take over once a critical threshold of tau aggregates is reached. This distinct-stage process also aligns with recent evidence of trans-neuronal tau spreading being critical for the initial expansion of tau pathology, but local replication of tau pathology driving accumulation of tau aggregates in later stages^59^. Stemming from this model, our work has potential implications for therapeutic approaches. We propose that targeting Aβ fibrils and soluble p-tau in early AD may be a promising strategy to slow the formation of tau aggregates, and thereby to preserve cognitive abilities for a longer period. Preliminary evidence from recent phase 2 and 3 clinical trials suggest that anti-Aβ therapies promptly reduce soluble p-tau concentrations in both CSF and plasma, reenforcing the close relationship between Aβ pathology and increases in extracellular p-tau levels^60-62^. Applying the same principles to anti-tau therapies (see^63^ for a review), we speculate that approaches reducing tau production and phosphorylation (e.g. antisense oligonucleotides, post-translational modification modulators, passive immunotherapies) would be most effective early on. On the other hand, in late stages of the disease, when local self-replication of tau aggregates is the predominant pathway that drives tau accumulation, targeting tau aggregates may exert stronger clinical benefits. As such, it is possible that therapeutics acting on tau aggregation inhibition and active clearance of aggregates might be more beneficial in the dementia stage. Our findings may be a starting point for future precision-medicine interventions that target the dominant pathway that determines tau aggregation, which are Aβ and soluble p-tau increases in early AD and tau aggregates in late AD.

**Figure 6.**
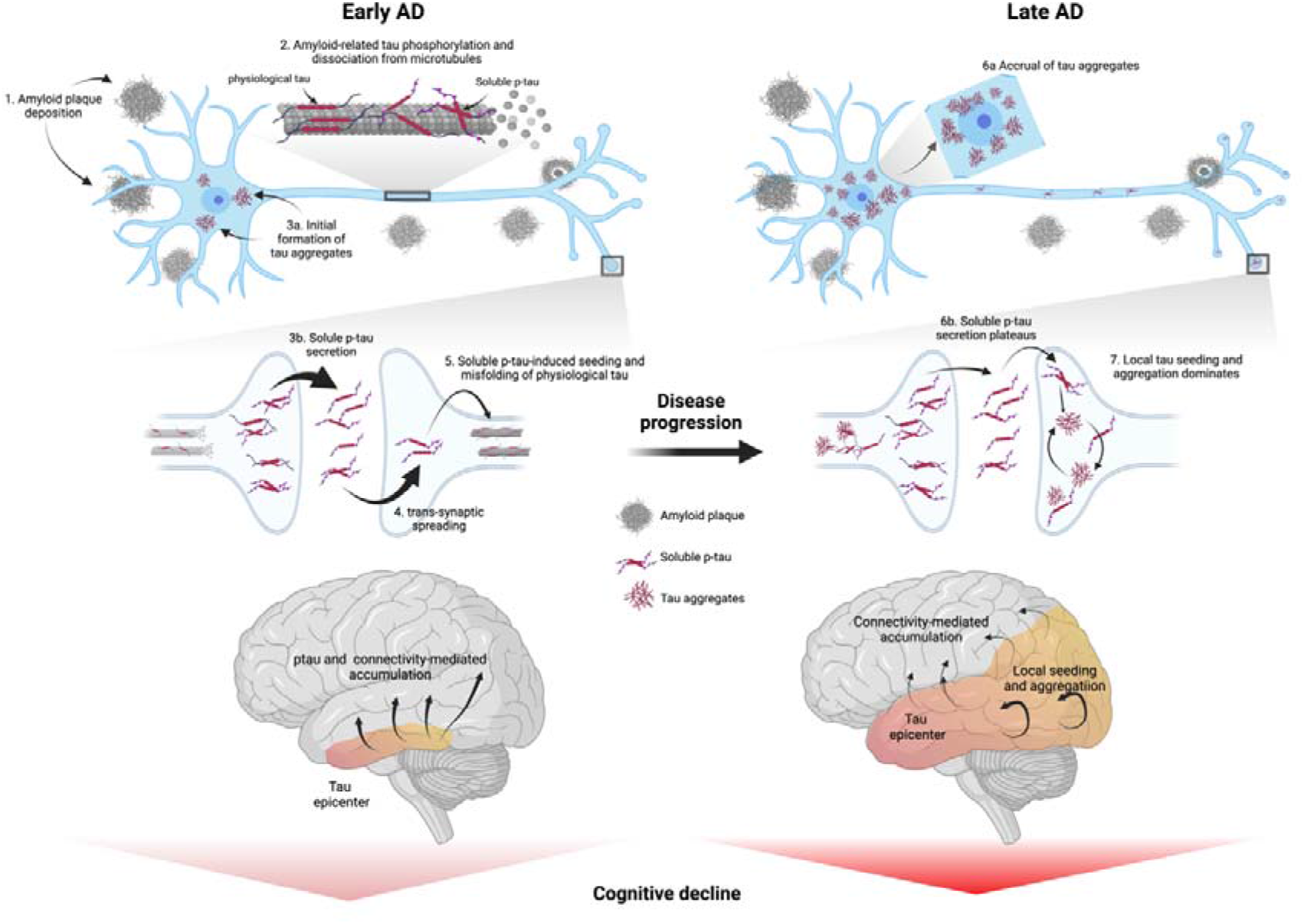
Proposed model of tau pathology accumulation in Alzheimer’s disease. In early AD, Aβ triggers increased concentrations and secretion of soluble p-tau, followed by post-synaptic uptake of p-tau seeds that lead to tau misfolding and aggregation. In late stages of the disease, when soluble p-tau concentrations have reached a plateau, local tau aggregates rather dominate in driving further local tau aggregation. Figure created with BioRender.com

## METHODS

### Participants - BioFINDER-2 cohort

The main cohort of interest included participants from the ongoing prospective Swedish BioFINDER-2 cohort (NCT03174938, http://www.biofinder.se/). All participants were recruited at Skåne University Hospital and the Hospital of Ängelholm, Sweden and the cohort covers the full spectrum of AD, ranging from adults with intact cognition or subjective cognitive decline, mild cognitive impairment (MCI), to dementia. The main inclusion criteria, as described previously^45^, were to be 40 years and older, being fluent in Swedish, having Mini-Mental State Examination (MMSE) between 27 and 30 for cognitively unimpaired (CU) participants, between 24 and 30 for MCI, and equal to or above 12 for AD dementia patients. Exclusion criteria were having significant unstable systemic illness, neurological or psychiatric illness, significant alcohol or substance misuse, or refusing lumbar puncture or neuroimaging. MCI diagnosis was established if participants performed below 1.5 standard deviation from norms on at least one cognitive domain from an extensive neuropsychological battery examining verbal fluency, episodic memory, visuospatial ability, and attention/executive domains. AD dementia diagnosis was determined on the criteria for dementia due to AD from the Diagnostic and Statistical Manual of Mental Disorders Fifth Edition and if positive on Aβ biomarkers based on the updated NIA-AA criteria for AD^64^. The study was approved by the Regional Ethics Committee in Lund, Sweden. All participants gave written informed consent to participate. All data for the current study was acquired between April 2017 and July 2021, and participants included needed to have CSF p-tau217 measurements and longitudinal tau-PET and cognitive data. After quality control of tau-PET (described below), the final sample included the following participants: 160 CU Aβ-negative, 51 CU Aβ-positive, 54 MCI Aβ-positive patients and 62 AD dementia patients. Follow-up time in tau-PET ranged from 0.73 to 2.5 years. CU participants are followed up every two years, while MCI and AD patients are seen annually.

### Participants - ADNI

ADNI is a multi-site study launched in 2003 as a public-private partnership. The primary goal of ADNI has been to test whether serial MRI, PET, other biological markers, and clinical and neuropsychological assessment can be combined to measure the progression of MCI and early Alzheimer’s disease. For up-to-date information, see www.adni-info.org. We selected participants who had longitudinal tau-PET, Aβ-PET (i.e. florbetaben or florbetapir) and CSF p-tau181 available in the ADNI database as of July 2021, which amounted to 122 participants out of the 777 individual participants with tau-PET. Out of those, only 3 were diagnosed with AD dementia and given the too small sample size, were not included in the study. The final sample of 119 thus included CU and MCI participants. Clinical status was assessed by ADNI: CU participants had a MMSE above 24, a Clinical Dementia Rating (CDR) score of 0, and were not depressed; participants with MCI had a MMSE above 24, a CDR of 0.5, objective memory-impairment based on education-adjusted Wechsler Memory Scale II and preserved activities of daily living.

### Image acquisition and processing - BioFINDER-2 cohort

MRI was performed using a Siemens 3T MAGNETOM Prisma scanner (Siemens Medical Solutions). Structural T1-weighted MRI images were acquired from a magnetization-prepared rapid gradient echo (MPRAGE) sequence with 1mm isotropic voxels. PET images were acquired on digital GE Discovery MI scanners. For tau-PET, acquisition was done 70-90 min post injection of ∼370 MBq [^18^F]RO948. For Aβ-PET, acquisition was done 90-110 min post injection of ∼185 MBq [^18^F]Flutemetamol. Images were processed according to our pipeline described previously^65^. Briefly, PET images were attenuation corrected, motion corrected, summed and registered to the closest T1-weighted MRI processed through the longitudinal pipeline of FreeSurfer version 6.0. Standardized uptake value ratio (SUVR) images were created using the inferior cerebellar gray matter as the reference region for [^18^F]RO948^66^, and the pons for [^18^F]Flutemetamol. PET SUVR images were then warped non-linearly to the Montreal Neurological Institute (MNI152) template space using Advanced Normalization Tools (ANTs)^67^ registration parameters of the T1-weighted MRI scan to the MNI152 template.

The cutoff for Aβ-positivity was 0.53 SUVR, defined from Gaussian mixture modeling (GMM) on a neocortical global Aβ region of interest (prefrontal, lateral temporal, parietal, anterior cingulate, and posterior cingulate/precuneus), as used previously^45^. This cutoff was used to classify CU and MCI participants, and a cutoff from CSF was used in AD dementia patients.

[^18^F]RO948 signal can be affected by off-target binding in the skull and meninges^37^. Given that the analyses focused on whole-brain cortical regions, we excluded a few participants with high levels of skull/meningeal binding, using a similar approach as described previously^58^. We calculated the SUVR ratio of a meningeal/skull mask to the average SUVR from GM, WM and CSF masks. Participants with a ratio above 1.75 were excluded, the latter having more than 1.75 times greater binding in the off-target region than in the brain. After such quality control, 10 CU Aβ-negative, 6 CU Aβ-positive and 2 MCI Aβ-positive participants with longitudinal tau-PET were excluded from the final sample.

### Image acquisition and processing - ADNI

Details on acquisition procedures of PET and MRI images in the ADNI cohort can be found elsewhere (http://adni.loni.usc.edu/methods/documents/). Briefly, structural MRI data was acquired on 3T scanners, using 3D T1-weighted MPRAGE sequences with 1mm isotropic voxel-size. PET data was acquired at standardized time-intervals post injection of ^18^F-labeled tracers. Flortaucipir was used for tau-PET and images were acquired 75-105 minutes post-injection. For Aβ-PET, florbetapir (image acquisition 50-70 minutes post-injection) and florbetaben (starting in ADNI3, image acquisition 90-110 minutes post-injection) were used. All image processing was done locally. Images were realigned, averaged, resliced to 1.5mm^3^, and smoothed to a resolution of 8mm^3^ FWHM. Each PET image was registered to the closest structural T1-weighted MRI image. SUVR images were created using the inferior cerebellum as the reference region for flortaucipir, and the whole cerebellum for florbetapir and florbetaben. Each T1-weighted image was also registered to the MNI template space using ANTs. Applying these registration parameters, SUVR PET images were then also registered to the MNI space for further analyses.

Aβ positivity was taken from the thresholds established by the ADNI PET core. The cutoff for positivity from a global composite cortical region referenced to the whole cerebellum was 1.11 SUVR for florbetapir and 1.08 for florbetaben^68^. Further, to be able to analyze all participants together despite two Aβ tracers being used, global Aβ SUVR were converted to the Centiloid scale^69^. As such, analyses with Aβ-PET in ADNI were done based on the global Aβ centiloid score and not at the regional level.

### Cerebrospinal fluid markers

In BioFINDER-2 CSF p-tau217 was measured using immunoassays on the Meso-Scale Discovery platform developed by Eli Lilly as described previously^39,70^. Dichotomization into CSF p-tau217 positivity was determined at a cutoff of 114.4 pg/ml, determined from GMM on measures from the whole BioFINDER-2 baseline data (n=943). Note that this threshold is virtually the same as when using 2 standard deviations from the mean of CU Aβ-negative participants (111.4 pg/ml). CSF Aβ42 and Aβ40 were measured using the Elecsys immunoassays (Roche Diagnostics)^16^. AD patients did not undergo Aβ-PET, and thus a pre-established cutoff of 0.08 on the CSF Aβ42/40 ratio was used to define Aβ-positivity^45^.

In ADNI CSF p-tau181 was measured at the University of Gothenburg using Elecsys immunoassays (Roche Diagnostics) and CSF p-tau217 was unavailable. CSF p-tau positivity was determined based using pre-established cut-offs at 21.8pg/ml^16^.

### Neuropsychological measures

In BioFINDER-2, cognitive decline was also investigated. We calculated a modified Preclinical Alzheimer’s Cognitive Composite (mPACC) as a composite measure to capture early cognitive decline^71,72^. The tests included in mPACC were the MMSE, ADAS-cog delayed recall, Trail-Making Test version A and Category Fluency of animals. The original PACC includes two measures of memory recall (Logical Memory and the Free and Cued Selective Reminding Test). Given that in our cohort only one memory score was available, ADAS-cog delayed recall was assigned a double weight to maintain the same proportion of memory as in the original composite score as done previously^73^. Further, the Digit Symbol Substitution Test used in the original PACC was replaced by Trail-Making Test A, to avoid missing values, as not all MCI patients completed the Symbol Digit Modalities Test. All tests were z-scored based on the mean and standard deviation of CU Aβ-negative participants over 50 years old, and then averaged to generate the mPACC score. Participants had two to four cognitive assessments.

To calculate change in cognition over time, linear mixed effect models with random slope and intercept were fitted with mPACC scores as the dependent variable and time in years from baseline as the independent variable. The slope of each participant from those models then represented rate of change in mPACC per year. The same approach was applied to MMSE scores only, to have an alternate measure of cognitive change over time.

### Regional measures and rate of change of PET SUVR

In both cohorts, all PET (Aβ and tau) images were parcellated into 200 regions corresponding to the widely used Schaefer functional atlas^74^. Average SUVR were extracted from these 200 parcels covering the entire neocortex, after masking each ROI with a grey matter mask to ensure that the final values to be used in analyses were least contaminated by binding from the white matter or CSF.

To calculate the rate of change in tau-PET over time, linear mixed effect models with random slope and intercept were fitted for each brain region, with tau-PET SUVR as the dependent variable and time in years from baseline as the independent variable. Participants had two or three tau-PET scans. The slope of each participant from those models then represented tau-PET SUVR change per year, referred to as rate of tau aggregates accumulation.

### Defining tau-PET epicenters

We defined tau-PET epicenters at the individual level from baseline tau-PET data, representing the regions with the highest probability of being abnormal using GMM. Given the skewed distribution of tau-PET, with many participants and many brain regions not exhibiting high levels of tau aggregates, we used GMM to separate target binding from non-specific binding. We fitted 2-distribution GMM on each brain region to separate both signals and extracted the probability to fall on the right-most distribution (“high” tau-PET distribution) for each participant^33,34,41^. Since this right-most distribution likely reflects abnormal tau-PET signal, the GMM probability represents the probabilistic measure of tau positivity. The GMMs were fitted on the whole BioFINDER-2 baseline tau-PET data (n=934), to have distributions corresponding as best as possible to the full range of tau-PET SUVR values across the AD spectrum. In ADNI, GMM were fitted on the sample with longitudinal tau-PET. A few parcels in the somatomotor regions with low tau-PET SUVR showed almost same fit between one or two distributions. They were excluded from the epicenter selection given that we could not clearly separate high-from low-tau distributions. This GMM probabilistic value was then multiplied with the tau-PET SUVR to obtain a SUVR score that was cleaned from unspecific signal. To define tau-PET epicenters at the individual level, the top 10 regions with the highest probability of SUVR weighted by GMM were selected and used for further connectivity-based analyses^33,40^.

### Functional connectivity template and analyses

To investigate if tau-PET accumulation was linked to functional brain architecture, we derived a template functional connectivity matrix from 69 CU from the ADNI cohort who were Aβ-negative and have low tau-PET binding (global SUVR<1.3). The steps to derive the template matrix have been described elsewhere (Franzmeier et al, preprint, 2021). Briefly, functional images were realigned to the first volume and co-registered to the native T1 images. Further processing included detrending, band-pass filtering (0.01-0.08Hz), and regressing out nuisance covariates (average white matter and CSF signal and motion parameters). Scrubbing of frames with frame displacement greater than 1 mm (along with the frame prior and the two subsequent frames) was applied and only participants with less than 30% of data had to be censored were retained. Functional connectivity matrices were created according to the functional Schaefer atlas of 200 parcels. Fisher-z correlations between time-series averaged across voxels within each ROI were determined to assess subject-specific functional connectivity matrices. All individual matrices were averaged and thresholded at 30% density. The average functional connectivity was then converted to a distance-based connectivity matrix, where then shorter path-length between ROIs represent stronger connectivity^75^. To link functional connectivity and tau aggregates accumulation, we calculated the distance-based functional connectivity of brain regions to the tau epicenters as defined above. Both at the group level and at the individual level, across all remaining brain region not an epicenter (n=190), we correlated tau-PET rate of change in each region to its connectivity-based distance to the tau epicenters. As such, we could measure the strength of the association between tau-PET accumulation and connectivity to tau epicenters across the whole brain, represented as a standardized β-value for each participant. Negative β-values were expected, meaning that stronger connectivity (represented as smaller values given that connectivity measures were converted to distance-based) would be associated with greater tau-PET change. From this connectivity-based analyses, for each participant, we then grouped the 190 non-epicenter regions into quartiles based on their connectivity to the tau epicenters (Q1 to Q4). The top 25% of regions with highest connectivity to the tau epicenters were part of Q1, the following 25% in Q2, etc. Average tau-PET rate of change in each quartile was calculated and used in further statistical analyses.

### Statistical analysis

All analyses were performed separately in the non-demented group (CU and MCI Aβ-positive) vs. the AD dementia group in BioFINDER-2. In ADNI analyses were restricted to the non-demented group. Importantly, given the smaller sample size in ADNI, only analyses with sufficient power (i.e., 80% and significance level of 0.05) based on effect sizes from BioFINDER-2 were performed in ADNI. The sample size estimates necessary to conduct each analysis in ADNI was calculated using the R package *pwr*. Details pertaining to the ADNI cohort are presented in the Extended Data. All analyses were performed in R version 4.0.5.

As a first step, we investigated the main factors related to accumulation of tau aggregates (measured as tau-PET rate of change), focusing on regional Aβ-PET SUVR and CSF p-tau217. In each of the 200 brain parcels, we applied different linear regression models, with tau-PET rate of change in each parcel as the dependent variable and age and sex as covariates. Models included progressively more dependent variables, starting with (1) regional Aβ-PET SUVR alone, (2) CSF p-tau217 alone, (3) regional Aβ-PET SUVR and CSF p-tau217, and (4) regional Aβ-PET SUVR, CSF p-tau217 and baseline regional tau-PET SUVR. We report results in regions where coefficients of regional Aβ-PET SUVR or CSF p-tau217 were considered significant, at a p-value < 0.05 after FDR correction. Based on the results from linear models, we also tested the mediating effect of CSF p-tau on regional Aβ-PET SUVR and regional tau-PET rate of change region-wise. Mediation analyses were conducted using the R package *mediation*. All paths of the mediation model were controlled for age and sex. Significance of the mediation effect was determined using 1000 bootstrapping iterations.

For each participant, linear models were fitted across all non-epicenter regions between connectivity to tau epicenters and tau-PET rate of change, resulting in a β-value. This β-value was then correlated with CSF p-tau concentrations, adjusting for age, sex and global Aβ-PET. Repeated measures ANOVA focusing on the interaction between CSF p-tau concentrations and rate of change in the different quartiles are also reported.

Focusing on the associations between tau measures and cognitive decline, we tested the mediating effect of (1) the β-value linking connectivity and tau aggregates accumulation across the brain and (2) tau-PET rate of change in Q1 on CSF p-tau and cognitive decline (measured either as slope of mPACC or slope of MMSE). Mediation models were performed as mentioned above. All paths of these mediation models were controlled for age, sex and years of education.

## Data Availability

Anonymized data from BioFINDER-2 (PI: OH) can be shared to qualified academic researchers after request for the purpose of replicating procedures and results presented in the study. Data transfer must be in agreement with EU legislation regarding general data protection regulation and decisions by the Ethical Review Board of Sweden and Region Skane, which should be regulated in a data transfer agreement. ADNI is a public access dataset and can be data can be obtained after application at http://adni.loni.usc.edu/. The template functional connectivity matrix created for the analyses can be available by contacting NF.

## ACKNOWLEDGMENTS

We would like to acknowledge all the BioFINDER team members as well as participants in the study and their family members for their dedication. Acknowledgement is also made to the donors of the Alzheimer’s Disease Research, a program of the BrightFocus Foundation, for support of this research (A2021013F). Work at the authors’ research center was supported by the Swedish Research Council (2016-00906), the Knut and Alice Wallenberg foundation (2017-0383), the Marianne and Marcus Wallenberg foundation (2015.0125), the Strategic Research Area MultiPark (Multidisciplinary Research in Parkinson’s disease) at Lund University, the Swedish Alzheimer Foundation (AF-939932), the Swedish Brain Foundation (FO2021-0293), The Parkinson foundation of Sweden (1280/20), the Konung Gustaf V:s och Drottning Victorias Frimurarestiftelse, the Skåne University Hospital Foundation (2020-O000028), Regionalt Forskningsstöd (2020-0314) and the Swedish federal government under the ALF agreement (2018-Projekt0279). The precursor of ^18^F-flutemetamol was sponsored by GE Healthcare. The precursor of ^18^F-RO948 was provided by Roche. The funding sources had no role in the design and conduct of the study; in the collection, analysis, interpretation of the data; or in the preparation, review, or approval of the manuscript.

Data collection and sharing for this project was funded by the Alzheimer’s Disease Neuroimaging Initiative (ADNI) (National Institutes of Health Grant U01 AG024904) and DOD ADNI (Department of Defense award number W81XWH-12-2-0012). ADNI is funded by the National Institute on Aging, the National Institute of Biomedical Imaging and Bioengineering, and through generous contributions from the following: AbbVie, Alzheimer’s Association; Alzheimer’s Drug Discovery Foundation; Araclon Biotech; BioClinica, Inc.; Biogen; Bristol-Myers Squibb Company; CereSpir, Inc.; Cogstate; Eisai Inc.; Elan Pharmaceuticals, Inc.; Eli Lilly and Company; EuroImmun; F. Hoffmann-La Roche Ltd and its affiliated company Genentech, Inc.; Fujirebio; GE Healthcare; IXICO Ltd.; Janssen Alzheimer Immunotherapy Research & Development, LLC.; Johnson & Johnson Pharmaceutical Research & Development LLC.; Lumosity; Lundbeck; Merck & Co., Inc.; Meso Scale Diagnostics, LLC.; NeuroRx Research; Neurotrack Technologies; Novartis Pharmaceuticals Corporation; Pfizer Inc.; Piramal Imaging; Servier; Takeda Pharmaceutical Company; and Transition Therapeutics. The Canadian Institutes of Health Research is providing funds to support ADNI clinical sites in Canada. Private sector contributions are facilitated by the Foundation for the National Institutes of Health (www.fnih.org). The grantee organization is the Northern California Institute for Research and Education, and the study is coordinated by the Alzheimer’s Therapeutic Research Institute at the University of Southern California. ADNI data are disseminated by the Laboratory for Neuro Imaging at the University of Southern California.

