## Supplementary material for "Amyloid-associated increases in soluble tau is a key driver in accumulation of tau aggregates and cognitive decline in early Alzheimer"

**Supplementary Results**

In the ADNI cohort, given the smaller sample size, only the results that could be validated with sufficient power (80% and alpha=0.05) were conducted. As such, the regional analyses relating soluble p-tau to local accumulation of tau aggregates were conducted, as well as the connectivity-based analyses on tau aggregates accumulation. Only 28 ADNI participants had longitudinal cognitive assessments, which was insufficient to test the mediations as done in BioFINDER-2. Based on effect sizes determined in BioFINDER-2, sample sizes between 75 and 82 participants would have been needed in order to assess effects of soluble p-tau on cognitive decline. Similarly, given the very few AD dementia patients with longitudinal tau-PET in ADNI (n=3), analyses pertaining to the dementia stage could not be conducted in this cohort.

In ADNI, as in BioFINDER-2, soluble p-tau was associated with regional accumulation of tau aggregates over time primarily in temporo-parietal regions, which remained significant when accounting for global Aβ-PET and baseline regional tau-PET SUVR (Extended Data Fig. 2). Given that two different Aβ-PET radiotracers were used in ADNI, global centiloid score was used as the Aβ measure. As such, the mediating effect of soluble p-tau on Aβ levels and rates of tau aggregates accumulation was done using global cortical measures. Overall, 30% (p=0.008) of the association between global Aβ and global accumulation of tau aggregates was mediated by soluble p-tau in Aβ-positive non-demented participants (Extended Data Fig. 2D). Further, all connectivity-based analyses were validated in ADNI (Extended Data Fig. 3). The overall connectivity-based association with tau-PET rate of change (β-value) was related to soluble p-tau levels (standardized coefficient=-0.236, p=0.012), and there was a significant interaction between soluble p-tau and connectivity-based tau aggregates accumulation by quartiles (p=0.014).

|  | **Aβ-negative controls**  **(n=52)** | **Aβ-positive non-demented**  **(n=67)** |
| --- | --- | --- |
| **Age (years)** | 71.7 (6.9) | 74.5 (6.7) |
| **Sex F (%F)** | 21 (40%) | 28 (42%) |
| **Education (years)** | 16.7 (2.4) | 16.6 (2.3) |
| ***APOE*ε4 carriers** (%) | 13 (25%) | 42 (63%) |
| **CSF p-tau181** (pg/ml) | 20.33 (6.98) | 33.54 (16.85) |
| **MMSE** | 29.3 (1.0) | 28.4 (1.9) |
| **Aβ Centiloid** | -3.8 (13.8) | 77.4 (33.6) |
| **tau-PET follow-up time** (years) | 2.3 (1.1) | 1.7 (0.6) |

**Extended Data Table 1: ADNI sample characteristics**
Data are presented as mean ± standard deviation unless specified otherwise.
Abbreviations: Aβ= beta-amyloid; *APOE*ε4= apolipoprotein E genotype (carrying at least one ε4 allele); CSF p-tau181= cerebrospinal fluid phosphorylated tau 181; MMSE= Mini-Mental State Evaluation; PET= positron emission tomography.

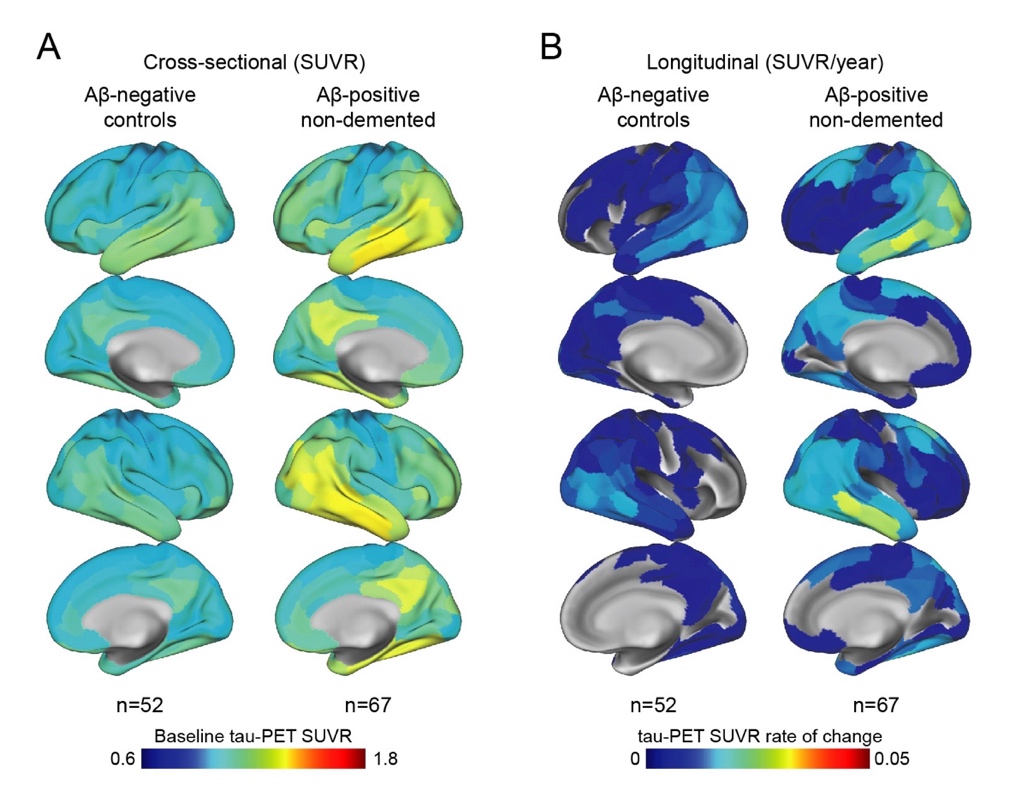

**Extended Data Figure 1. Mean spatial distribution of cross-sectional flortaucipir tau-PET SUVR and longitudinal rate of change in ADNI**

(A) Surface renderings of average baseline tau-PET SUVR in Aβ-negative controls and Aβ-positive non-demented participants in the 200 parcels from the Schaefer 200-ROI atlas (B) Surface renderings of yearly tau-PET SUVR rate of change derived as the slope from linear mixed-effect models in the same participants group as in (A)

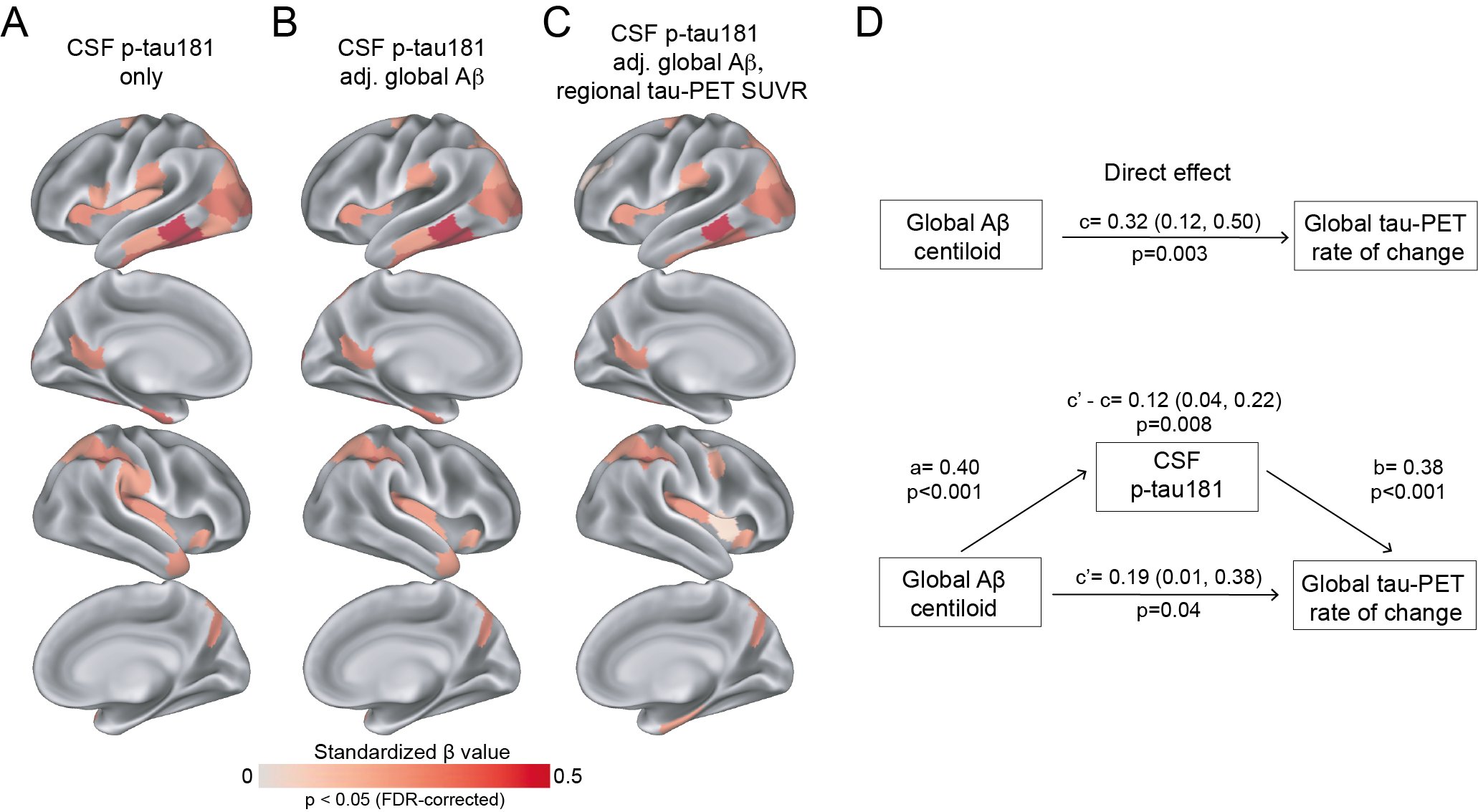

**Extended Data Figure 2. CSF p-tau181 and global Aβ associations with regional tau aggregates accumulation in Aβ-positive non-demented participants in ADNI**

(A) Standardized beta coefficient of CSF p-tau181 in regions where CSF p-tau181 relates to regional tau-PET rate of change, adjusting for age and sex (B) Standardized beta coefficient of CSF p-tau181 in regions where CSF p-tau181 relates to regional tau-PET rate of change, when adjusting for global Aβ SUVR, age and sex, and (C) when also adjusting for regional baseline tau-PET SUVR (D) Mediating effect of CSF p-tau181 on global Aβ SUVR accumulation of tau aggregates across the brain (average rate of change across 200 regions). All regions shown on the brain are significant at p<0.05 after FDR-correction.

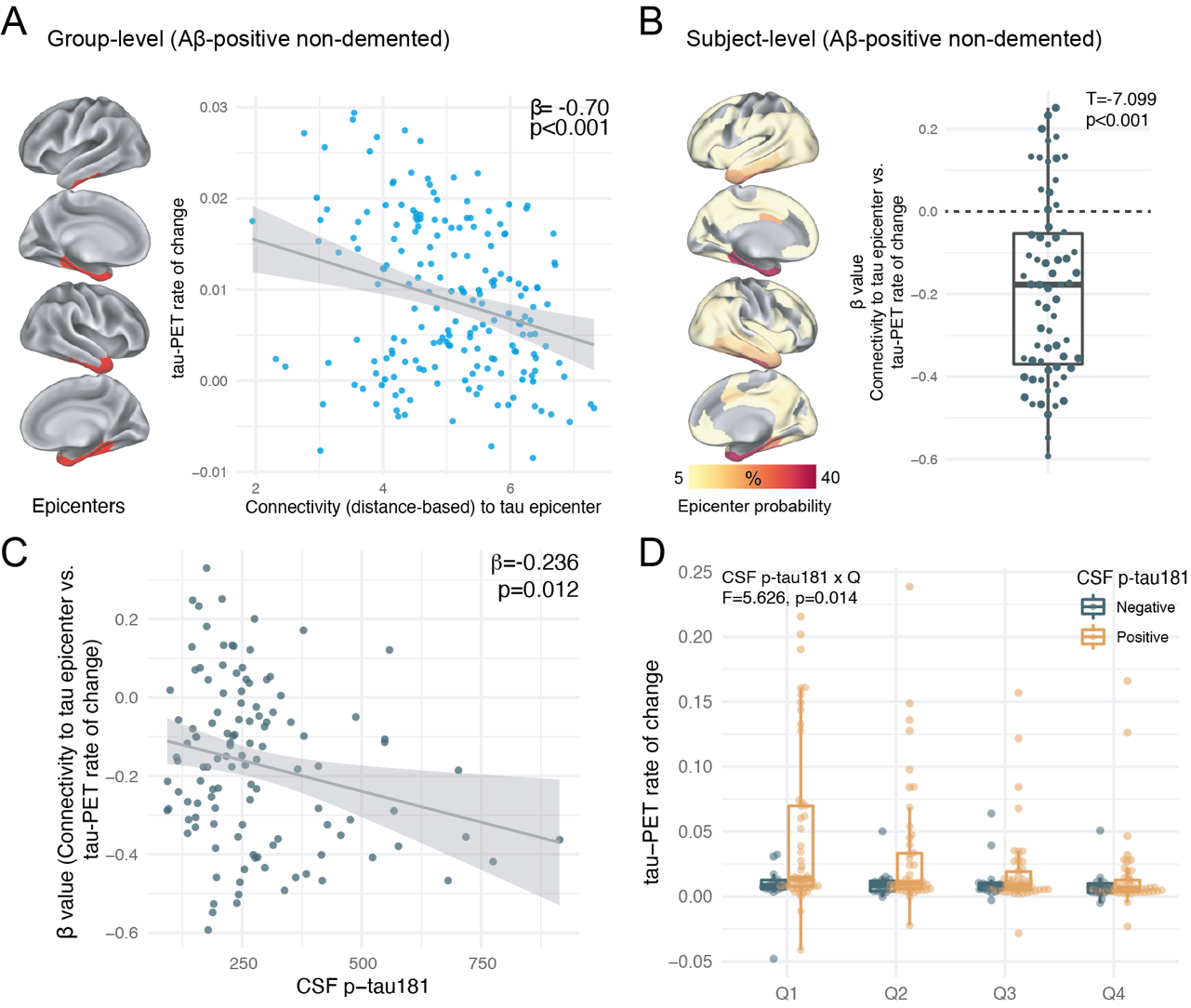

**Extended Data Figure 3. Individualized connectivity-based associations of tau-PET rate of change over time and CSF p-tau181 in Aβ-positive non-demented participants in ADNI**

(A) Group-level analysis showing how connectivity to the tau epicenters (projected on the glass brains) relates to tau-PET rate of change across the whole brain. Each dot represents a brain region. Regions more strongly functionally connected to the epicenters have greater rate of tau-PET accumulation (B) Repeating the same approach depicted in (A) at the individual level, the values on the glass brains represent the percentage that each region is classified as an epicenter. The box plot shows the individual β-value from the correlation between tau-PET rate of change and connectivity-based distance to epicenters across all brain regions (C) Scatter plot of the associations between CSF p-tau181 and the β-values of epicenter connectivity to tau-PET rate of change. The expected negative association suggests that higher CSF p-tau181 is associated with the overall pattern of tau-PET change in more functionally connected regions to epicenters (D) Average tau-PET rate of change in regions split into quartiles based each region’s connectivity to the tau epicenters (Q1 represents top 25% regions with strongest functional connectivity to the epicenters, etc.). In each quartile the dots represent each participant.
